# Clustering of shoulder movement patterns using K-means algorithm based on the shoulder range of motion

**DOI:** 10.1101/2023.07.31.23293406

**Authors:** Jun-hee Kim

## Abstract

**Context:** Categorization in medicine is used to enhance understanding of a disease or syndrome and apply it to treatment and is based on human clinical experience or theory. Cluster analysis using the K-means algorithm is an unsupervised machine learning method that classifies clusters based on numerical data. The purpose of this study was to classify subjects into clusters using K-means algorithm based on shoulder range of motion (ROM) and identify the characteristics of the clusters.

**Design:** Cross-sectional study

**Methods:** 551 data samples measured in the 5th Size Korea Anthropometric Survey (2003∼2004) were used. Clustering was performed using the K-means algorithm, and the appropriate number of clusters was determined using the elbow curve and silhouette score. The characteristics of the clusters were analyzed by comparing the average values of shoulder ROM in the clusters.

**Results:** The appropriate number of classifications of clusters according to the shoulder ROM was 8. Clusters 1 and 5 had the lowest flexion range, and clusters 7 and 8 had low internal rotation and shoulder horizontal adduction ranges. Clusters 2 and 6 exhibited the highest flexion and overall high flexibility. Clusters 3 and 4 showed moderate flexion ranges but low horizontal adduction ranges. Shoulder movement patterns were classified into a total of 8 clusters according to the shoulder ROM.

**Conclusion:** Based on this clustering system, it was possible to identify the pattern of shoulder movement in ordinary people, and it could be used as basic data to identify and treat diseases or syndromes according to the pattern.

## Introduction

Categorization is a broad concept that classifies or groups objects, concepts, social groups, etc. according to common characteristics.^1^ In other words, categorization means the act of distinguishing between things with different properties and collecting things with the same properties.^1^ Categorization has a deep relationship with human cognitive ability, and through categorization, humans have been equipped with advanced thinking abilities such as observation, comparison, understanding, reasoning, judgment, and application.^1, 2^

Categorization is one of the important concepts in the field of medicine. Pathology is a representative academic field that uses the concept of categorization the most in medicine.^3^ Pathology is a discipline that systematically analyzes and organizes the changes that occur in the human body according to diseases.^3^ As numerous diseases have been classified and analyzed systematically, human health has improved steadily over a long period of time.^4, 5^ Depending on the categorization of the disease, it has been used to find the cause of the disease, select the treatment according to the disease, and predict the patient’s prognosis.^5–7^

In addition to categorization of diseases, patients are categorized into different syndromes to classify people who experience a set of symptoms and signs into a specific set.^8^ Regarding musculoskeletal disorders, various syndromes are classified according to their symptoms and signs, and treatments are systematically used to recover these classified syndromes.^8–11^ Janda proposed a “crossed syndrome” of muscle imbalance in 1979 based on her clinical observations and studies, and theorized that this physical syndrome due to muscle imbalance was predictable and related to the entire motor system.^12^ According to his theory, upper crossed syndrome was defined as abnormal alignment and pain due to overactivity of the upper trapezius, levator scapulae, sternocleidomastoid, and pectoralis major muscles, and inhibition of the deep cervical flexor, lower trapezius, and serratus anterior.^12^ Since the imbalance of these muscles leads to motor dysfunction, it is said that resolving this imbalance improves shoulder function and relieves pain.^12^ Sahrmann (2002) defined movement impairment syndrome as a condition in which pain or functional impairment occurs due to imbalance of muscles caused by incorrect posture or repeated movements.^13^ Based on her clinical experiences and studies, shoulder movement impairment syndromes can be classified into 8 types according to the alignment of the shoulder joints or movement patterns.^13^ It is said that pain, abnormalities and functional limitations of the shoulder joints are restored by restoring the imbalance of muscles or correcting the faulty movements of each syndrome with precise movements.^13^

Clustering analysis is an old data analysis technique first used in psychology by Driver and Kroeber in 1932, but has been rapidly systematized in recent years with advances in artificial intelligence and machine learning.^14^ Clustering is a similar concept to categorization, but clustering analysis is a way of classifying objects into clusters by unsupervised machine learning algorithms.^14, 15^ It is an analysis technique that identifies differences between clusters by measuring the similarity of each piece of data and categorizing them into multiple clusters.^14, 15^ Clustering enables a process of grouping objects into clusters based on characteristics or similarities inherent in objects only with quantified data without subjective human judgment or bias.^14, 15^ The K-means algorithm is one of the unsupervised learning algorithms used for clustering and is a method of grouping data into a predefined number of clusters.^16–19^ Since the K-means algorithm is a distance-based algorithm, it is suitable for continuous data and can be applied to large-scale datasets because it is relatively simple, fast, and scalable.^16, 17, 20^

So far, in relation to physical health, subjects have been categorized according to diseases or syndromes based on characteristics such as movement patterns or muscle imbalances. In other words, the movement patterns were compared or analyzed by classifying only those with or without physical problems. Also, these categorizations have been theorized based on human observations and studies. However, the categorization of human movement patterns in which diseases or syndromes are not considered has not yet been studied. In addition, there has been no study that has categorized movement patterns through a clustering technique, one of unsupervised machine learning, based on physical data related to movement of normal adults. Therefore, the purpose of this study was to classify shoulder movement patterns using K-means clustering techniques based on the shoulder range of motion (ROM).

## Materials and methods

### 1. Data Source

This study used data samples obtained from the 5th anthropometric survey (2003-2004) of the xxxx xxxxxx (xxxxxx anthropometric survey) project of the xxxxxx Agency for Technology and Standards. Among a total of 551 data samples, 10 data samples with missing shoulder ROM were excluded, and 541 samples were finally selected. (Figure 1). The data samples included age and gender information, excluding personally identifiable information, and included shoulder flexion, extension, horizontal adduction, horizontal abduction, internal rotation, and external rotation ROM (Table 1). This study was approved by the Public Institutional Review Board Designated by Ministry of Health and Welfare (approval number: xxx-xxxxxx-xx-xxx)

**Figure 1.**
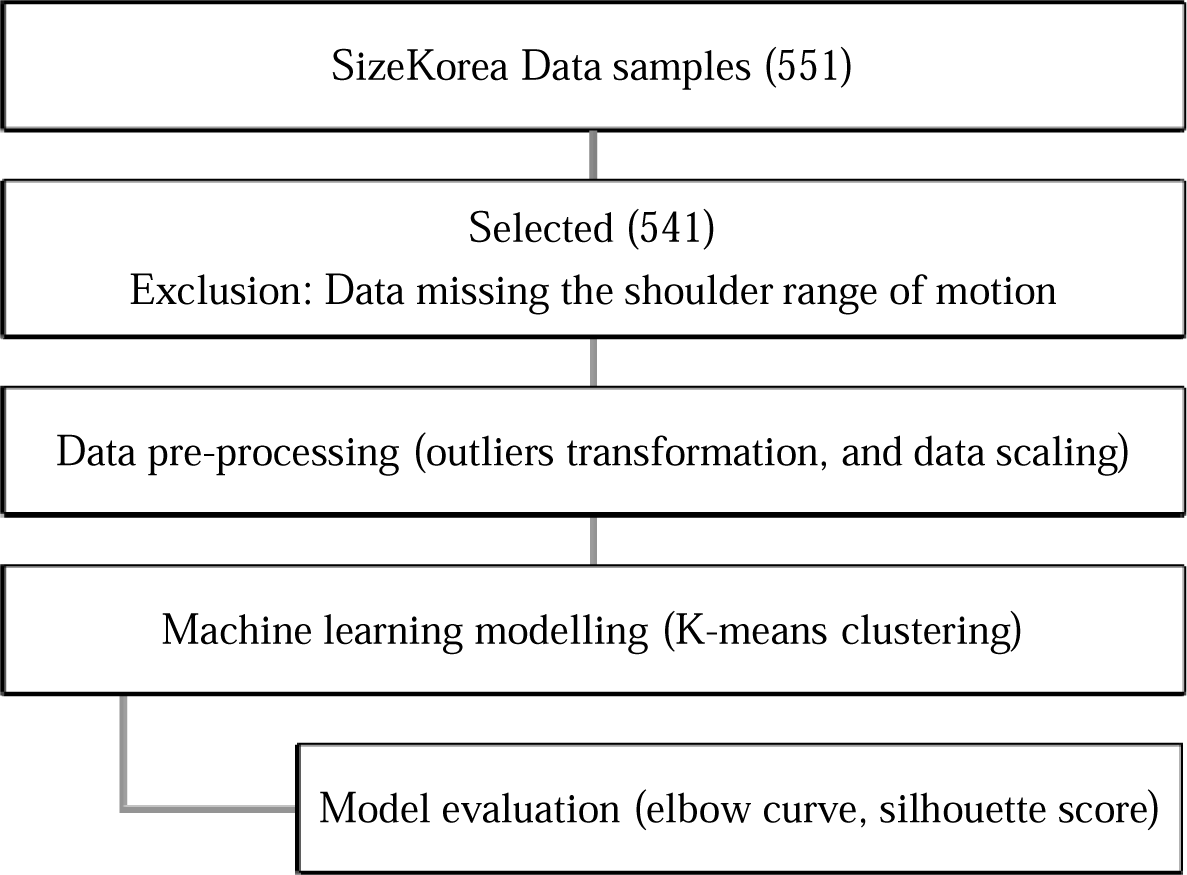
Flowchart of the study.

**Table 1.** Variables description.

| Variables | Description | Value |
| --- | --- | --- |
| age | Age | $24.34 \pm 5.53$ |
| sex | Gender | Male: 325, Female: 216 |
| flex | Shoulder flexion | $176.37 \pm 29.23$ |
| ext | Shoulder extension | $60.49 \pm 13.12$ |
| hor_add | Shoulder horizontal adduction | $53.72 \pm 11.36$ |
| hor_abd | Shoulder horizontal abduction | $117.18 \pm 14.32$ |
| int_rot | Shoulder internal rotation | $48.31 \pm 9.40$ |
| ext_rot | Shoulder external rotation | $64.32 \pm 10.48$ |

### 2. Data Preprocessing

Interquartile range (IQR) method was used to detect and convert outliers. Outliers outside the 1.5 * IQR point were detected at the 25% (Q1) and 75% (Q3) points in the data distribution, and the outlier data were converted to fit the 1.5 IQR point.^21^ Shoulder ROM data, which are continuous variables, were standardized using the StandardScaler function.^22^

### 3. Machine learning algorithms

The shoulder movement pattern clustering model was developed as a clustering technique using the K-means algorithm, an unsupervised machine learning technique. Since the K-means clustering technique requires pre-determination of the number of categorizations before model construction, the number of categorizations was set to be between 2 and 12.

### 4. Model evaluation

The elbow curve and silhouette score were used to select the number of shoulder movement patterns and evaluate the quality of the model. An elbow curve graphs the change in sum of squared within-cluster variation (SSW) according to the number of clusters (k). As the number of clusters increases, the SSW value decreases, and then the appropriate number of clusters is determined through an “elbow” point where the slope of the decrease becomes gentle.^23^ In addition, the silhouette score is calculated considering the similarity within the cluster of each data point and the distance between other clusters. The silhouette score ranges from -1 to 1, and the closer to 1, the better the classification of clusters, and the closer to 0, the less certain the clustering result.^24^

## Results

### 1. The number of clusters

In the shoulder movement pattern clustering model using the K-means algorithm, an elbow point did not appear according to the change in the number of clusters (Figure 2). In other words, the decreasing slope of SSW according to the cluster increase was relatively constant, and it was not possible to select the number of clusters using the elbow curve. However, the silhouette score was highest when the number of clusters was 8.

**Figure 2.**
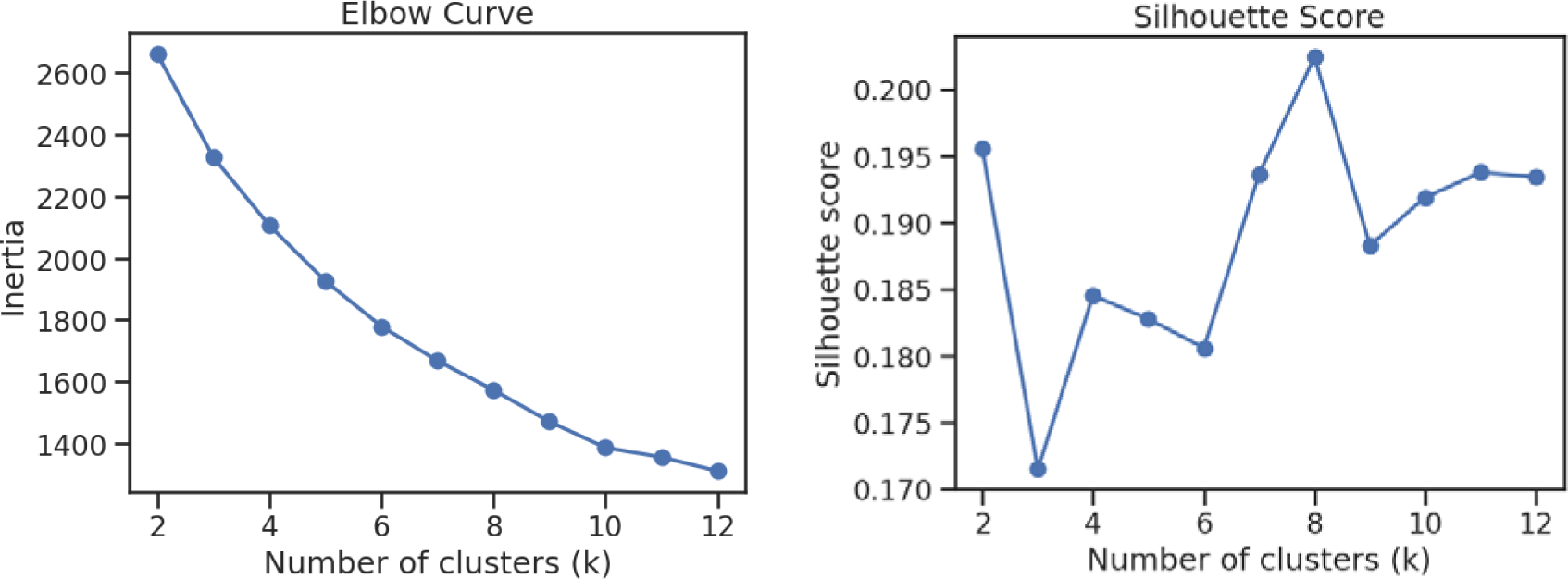
Elbow curve and silhouette score.

### 2. Clusters of shoulder movement pattern

Table 2 and Figure 3 show the 8 shoulder movement pattern clusters identified by the k-means algorithm clustering, their gender and age, and the ranges of motion for shoulder joint. Subjects with the movement pattern of cluster 1 had a relatively low range of external rotation and flexion compared to other clusters, but a high range of internal rotation. Subjects with the movement pattern of cluster 2 had the highest flexion range and relatively low horizontal abduction range compared to other clusters. Subjects with movement pattern of cluster 3 had higher rotation and horizontal abduction ranges than other clusters, but the rest ranges were not particularly high or low. Subjects with movement patterns of cluster 4 had a higher extension range than other clusters, but the rest ranges were not particularly high or low. Subjects with movement patterns of cluster 5 had higher ranges of extension and external rotation, but lower ranges of flexion and horizontal abduction than other clusters. Subjects with the movement pattern of cluster 6 showed the second highest flexion ROM, and horizontal adduction and abduction showed the highest ROM. Subjects with the movement pattern of cluster 7 showed the second lowest ROM for internal rotation and showed lower ranges of extension, horizontal adduction, and horizontal abduction than the other clusters. Subjects with movement pattern of cluster 8 showed the lowest internal rotation range compared to the other clusters.

**Table 2.** Shoulder movement pattern clusters and ranges of motion.

| Clusters | Gender<br>(M/F) | Age | Flexion | Extension | Internal<br>rotation | External<br>rotation | Horizontal<br>adduction | Horizontal<br>abduction |
| --- | --- | --- | --- | --- | --- | --- | --- | --- |
| Cluster 1 | 15/17 | 22.94 ± 5.34 | 143.56 ± 21.22 | 51.69 ± 8.83 | 55.16 ± 6.19 | 49.56 ± 4.59 | 56.38 ± 9.79 | 118.59 ± 13.68 |
| Cluster 2 | 97/43 | 24.37 ± 4.91 | 202.81 ± 10.37 | 61.37 ± 7.54 | 48.17 ± 4.17 | 65.75 ± 4.90 | 58.06 ± 5.37 | 107.99 ± 6.77 |
| Cluster 3 | 19/29 | 24.32 ± 6.38 | 170.50 ± 24.32 | 51.82 ± 8.73 | 53.58 ± 7.24 | 70.15 ± 6.42 | 45.88 ± 8.62 | 131.85 ± 10.58 |
| Cluster 4 | 36/29 | 25.02 ± 6.38 | 163.22 ± 25.02 | 68.15 ± 7.52 | 51.40 ± 4.94 | 57.08 ± 6.90 | 42.15 ± 7.67 | 118.37 ± 9.20 |
| Cluster 5 | 17/33 | 24.38 ± 5.33 | 143.46 ± 24.38 | 66.49 ± 12.19 | 52.74 ± 6.51 | 74.49 ± 5.79 | 56.00 ± 9.39 | 107.04 ± 14.26 |
| Cluster 6 | 65/29 | 24.01 ± 5.79 | 191.90 ± 24.01 | 66.04 ± 8.96 | 47.23 ± 5.64 | 68.73 ± 4.78 | 64.06 ± 7.01 | 134.85 ± 5.55 |
| Cluster 7 | 23/11 | 24.81 ± 5.34 | 163.82 ± 24.81 | 42.16 ± 5.22 | 44.38 ± 7.63 | 56.49 ± 7.27 | 34.88 ± 6.55 | 111.50 ± 8.05 |
| Cluster 8 | 53/25 | 24.50 ± 5.31 | 164.77 ± 24.50 | 59.10 ± 6.88 | 36.86 ± 4.57 | 63.60 ± 4.68 | 53.23 ± 9.05 | 111.09 ± 11.15 |

**Figure 3.**
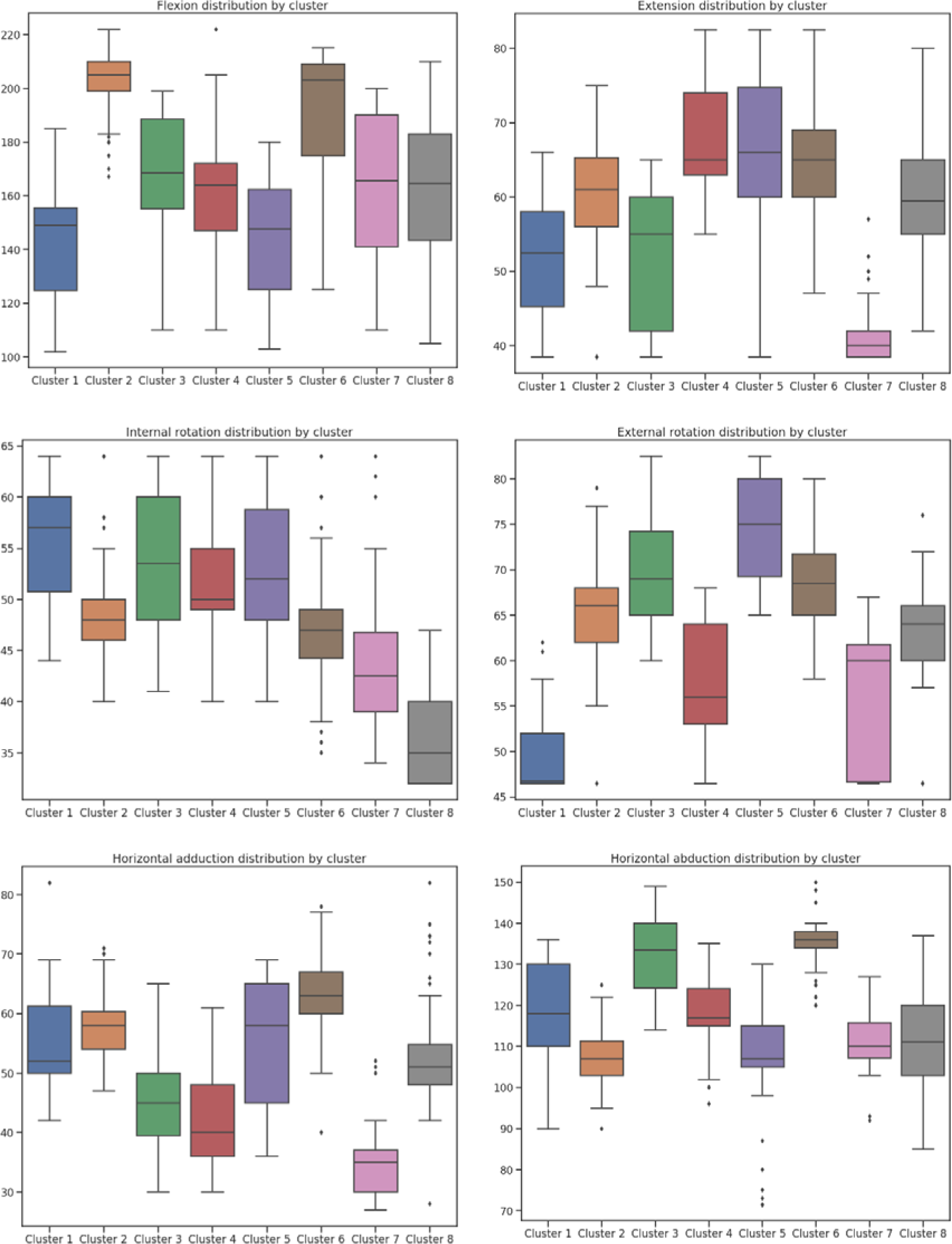
Shoulder movement pattern clusters and ranges of motion.

## Discussion

The purpose of this study is to perform clustering according to shoulder ROM with the K-means algorithm, an unsupervised machine learning data clustering algorithm that operates without human prior knowledge or prejudice. As a result of classification using the K-means algorithm, subjects formed a total of 8 clusters according to shoulder ROM, and each cluster showed different shoulder movement patterns.

Biomechanics is a study that analyzes the scientific principles related to the movement of the human body, and investigates and studies forces, alignments, and movements acting on the human body. Based on biomechanics, it is emphasized that incorrect postures or movements occurring in the human body can cause musculoskeletal disorders or syndromes and improving them can be a fundamental treatment for musculoskeletal related problems. Representatively, Sahrmann (2002) classified and defined eight types of syndromes based on the alignment of the scapulae or humerus that make up the shoulder girdle or the movement of joints such as the scapulothoracic or glenohumeral joints based on biomechanics.^13^ According to each shoulder movement impairment syndrome, abnormal shoulder movements were observed along with shortening of the length of specific muscles or weakness of specific muscles.^13^ Similar to the results of this study, the clustering analysis according to the shoulder ROM revealed 8 types of shoulder movement pattern clusters.

Sahrmann (2002) suggested that the shoulder flexion motion could be restricted in common in the 8 types of shoulder movement impairment syndrome.^13^ It has been reported that the flexion ROM is significantly reduced due to symptoms such as shoulder impingement accompanied by pain, insufficient scapular movement, or abnormal scapular movement.^13^ In addition to scapular movements, excessive abnormal gliding motion occurring in the glenohumeral joint can also affect the shoulder flexion ROM.^13^ In this study, clusters 1 and 5 are classified as having limited shoulder flexion angle. However, the difference between cluster 1 and cluster 5 was only the difference in the range of external rotation of the shoulder. Cluster 1 had a lower external rotation range compared to cluster 5. The pattern of limited shoulder flexion and external rotation suggests that cluster 1 probably exhibits a movement pattern similar to shoulder medial rotation syndrome in movement impairment syndrome.^13^ Camp (2017) found that reduced shoulder flexion and external rotation ROM increased the risk of elbow injury by 9% and 7%, respectively, in the study conducted on 132 adult pitchers.^25^ Based on the above study results, Cluster 1 may be at risk of elbow injury due to limitations in the ROM for shoulder flexion and external rotation. According to previous studies on shoulder impingement syndrome, also referred to as subacromial pain syndrome, and internal rotation ROM, it was reported that patients with this syndrome had a significantly reduced internal rotation ROM.^26–28^ Cluster 8 showed the lowest internal rotation ROM, although it did not seem to show much difference from the other clusters in other shoulder ranges of motion. Based on above study results, subjects who formed cluster 8 are likely suffering from or related to subacromial pain syndrome. In addition, it has been reported that a decrease in shoulder internal rotation angle is related to a decrease in horizontal adduction.^29^ This reduction in internal rotation and horizontal adduction can lead to abnormal humerus motion and reduced subacromial space, leading to subacromial pain syndrome as mentioned above.^29, 30^ Cluster 7 exhibited the second lowest internal rotation as well as the lowest horizontal adduction range. Therefore, compared to cluster 8, the movement pattern of cluster 7 was classified as a different cluster, but it can be assumed that there is a condition or risk of subacromial pain syndrome as well.

Clusters 2 and 6 showed the highest flexion range. However, cluster 6 showed a high range of both horizontal adduction and abduction compared to other clusters, while cluster 2 showed a medium range. In terms of the range of rotations, both clusters 2 and 6 showed intermediate ranges. Nonetheless, according to the above study, these two clusters would be unrestricted and flexible with no problems in their ROM and would have a much lower risk of shoulder musculoskeletal disorders than the other clusters. Clusters 3 and 4 have lower ranges of flexion, which is clinically important motion, than clusters 2 and 6, but still have a greater range than the other clusters. Also, the rotation ranges were high to medium. Nevertheless, since the range of shoulder horizontal adduction was the second and third lowest, respectively, there is a possibility that the risk of subacromial pain syndrome is higher than that of the clusters 2 and 6.

There are several limitations in this study. First, information such as disease or syndrome was not labeled in the data used for machine learning modeling in this study. Nevertheless, the original purpose of data collection used in this paper was to investigate standard human data, and the average age of the subjects was in their 20s, and the frequency of shoulder-related diseases would have been relatively low. Therefore, there would be no major errors in the clustering model through this study. Second, in this study, clusters were classified by differences in shoulder ROM values using the K-means algorithm, but these differences were not performed by a parametric test assuming a normal distribution. Therefore, it could not be shown that the difference in shoulder movement by cluster mentioned in this study was a significant difference by parametric test. Third, although movement patterns were classified into 8 clusters based on shoulder ROM data, it may not be clear to make a clinical interpretation of each cluster only by classifying shoulder ROM. In the future, further studies will be needed to determine whether clusters with these movement patterns have a causal relationship or association with specific shoulder diseases or syndromes.

## Conclusion

In this study, clustering analysis was performed using the K-means algorithm based on shoulder ROM data of flexion, extension, internal rotation, external rotation, horizontal adduction, and horizontal abduction. Shoulder movement patterns were classified into a total of 8 clusters according to the shoulder ROM. Clusters 1 and 5 showed a limited range of flexion common in movement impairment syndrome, as well as cluster 1 showed a limited range of external rotation. Clusters 7 and 8 were classified as having a reduced range of internal rotation, which is closely related to subacromial pain syndrome. In addition, cluster 7 showed the lowest range of horizontal adduction, which is another cause of subacromial pain syndrome. Clusters 2 and 6 showed high flexibility in the flexion angle, and the rest of the ROM also showed a medium to high level. Clusters 3 and 4 showed moderate range of flexion, but the range of shoulder horizontal adduction, which was associated with the risk of subacromial pain syndrome, was the second and third lowest, respectively.

## Data Availability

This study used data samples obtained from the 5th anthropometric survey (2003-2004) of the Size Korea (Korean anthropometric survey) project of the Korean Agency for Technology and Standards.

https://sizekorea.kr/

## Acknowledgments

This research did not receive any specific grant from funding agencies in the public, commercial, or not-for-profit sectors. The author’s responsibilities were as follows: xxx designed the study and wrote the manuscript; xxx analyzed the data; xxx edited the manuscript; xxx had primary responsibility for the final content of the manuscript; All authors read and approved the final manuscript.

